# Asymptomatic SARS-CoV-2 infection by age: A systematic review and meta-analysis

**DOI:** 10.1101/2022.05.05.22274697

**Authors:** Bing Wang, Prabha Andraweera, Salenna Elliott, Hassen Mohammed, Zohra Lassi, Ashley Twigger, Chloe Borgas, Shehani Gunasekera, Shamez Ladhani, Helen Siobhan Marshall

**Author notes:** Correspondence; Address: Women’s and Children’s Hospital, North Adelaide, 5006, South Australia, Australia.

## Abstract

**Objectives:** This systematic review and meta-analysis aimed to estimate the age-specific proportion of asymptomatic SARS-CoV-2 infected persons by year of age.

**Methods:** We searched PubMed, Embase, medRxiv and Google Scholar on 10 September 2020 and 1 March 2021. We included studies conducted during January to October 2020, prior to routine vaccination against COVID-19. Since we expected the relationship between the asymptomatic proportion and age to be non-linear, multilevel mixed-effects logistic regression (QR decomposition) with a restricted cubic spline was used to model asymptomatic proportions as a function of age.

**Results:** A total of 38 studies were included in the meta-analysis. In total, 6556 out of 14850 cases were reported as asymptomatic. The overall estimate of the proportion of people who became infected with SARS-CoV-2 and remained asymptomatic throughout infection was 44.1% (6556/14850, 95%CI 43.3%-45.0%). The asymptomatic proportion peaked in adolescents (36.2%, 95%CI 26.0%-46.5%) at 13.5 years, gradually decreased by age and was lowest at 90.5 years of age (8.1%, 95%CI 3.4%-12.7%).

**Conclusions:** Given the high rates of asymptomatic carriage in adolescents and young adults and their active role in virus transmission in the community, heightened vigilance and public health strategies are needed among these individuals to prevent disease transmission.

## 1. Introduction

The severe acute respiratory syndrome coronavirus 2 (SARS-CoV-2) pandemic causing coronavirus disease 2019 (COVID-19) has had a profound impact on public health, our daily life, and economies around the world. Asymptomatic infections have raised concerns about public health policies for managing epidemics because they are a potential source of transmission of the virus and a challenge for controlling the pandemic.^1,2^ A few systematic reviews have been conducted to determine the contribution of asymptomatic infection to SARS-CoV-2 transmission.^1,3-19^ Previous researchers attempted to synthesize the best available evidence in different age groups such as children, adults, and elderly.^5,7,11,13,16^ None, however, have investigated the proportion of asymptomatic SARS-CoV-2 infections throughout the course of infection by age. This review, therefore, aims to 1) identify, assess, and synthesize the evidence on the proportion of people infected with SARS-CoV-2 who were asymptomatic throughout the course of infection, and 2) to estimate asymptomatic proportion by age.

## 2. Methods

### 2.1. Study Design

We conducted this systematic review and meta-analysis based on the statement of the Preferred Reporting Items for Systematic Review and Meta-Analysis (PRISMA) guidelines^20^ (Figure 1). The study protocol was registered in the International Prospective Register for Systematic Reviews (PROSPERO, registration number: CRD42020209419).

**Figure 1.**
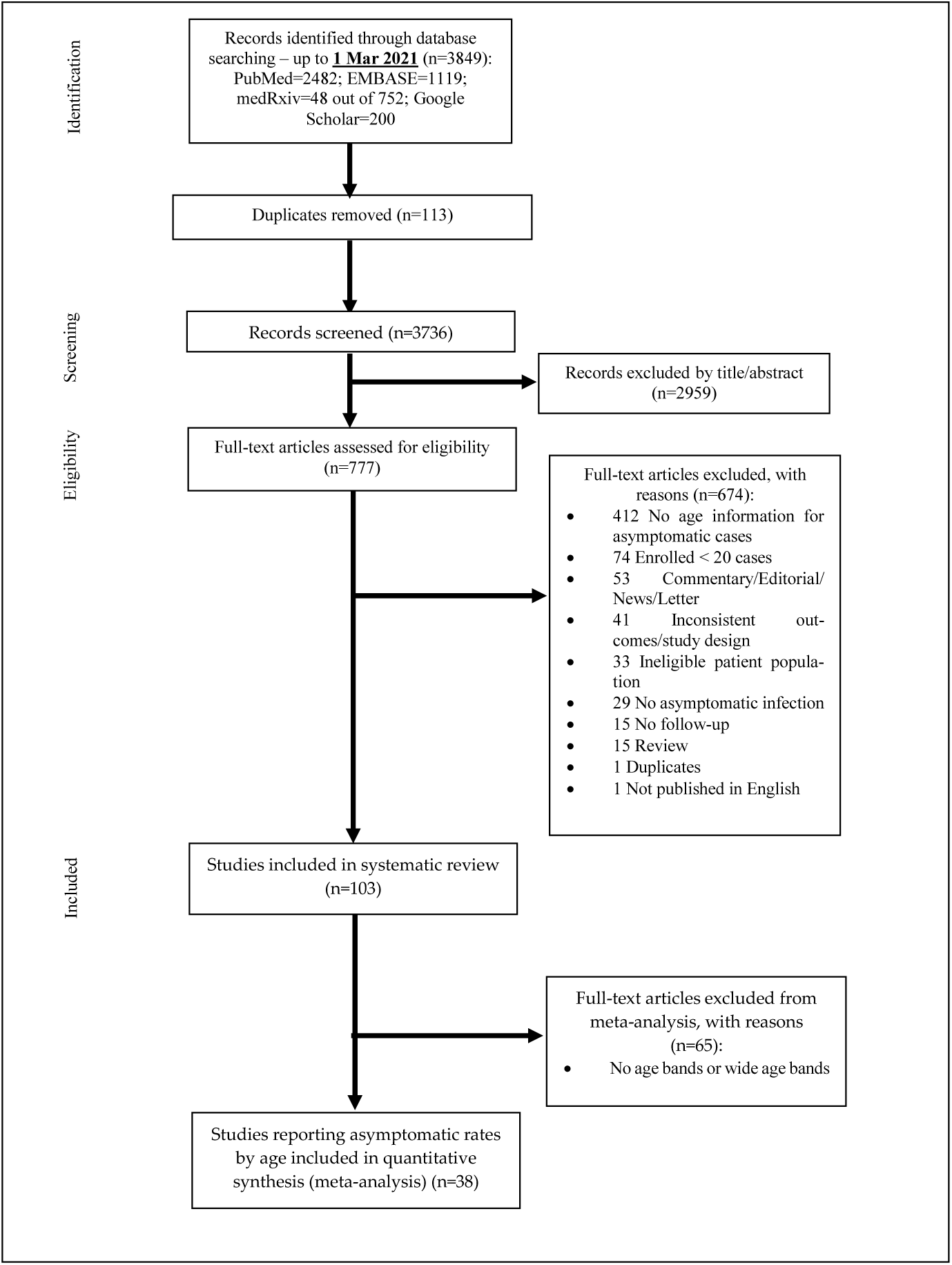
The Preferred Reporting Items for Systematic Reviews and Meta-analyses (PRISMA) flow diagram for article inclusion and exclusion

**Fig 2.**
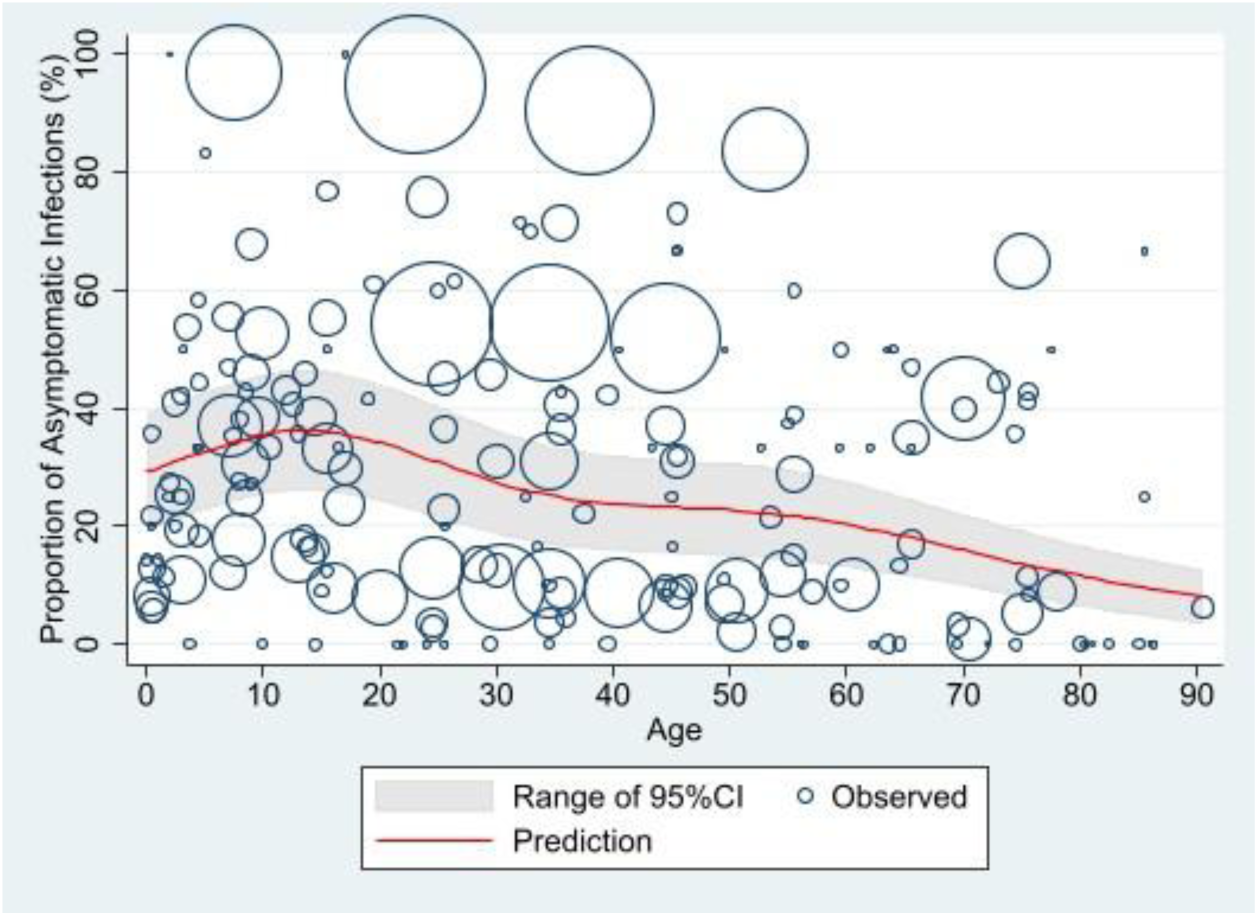
Estimates of asymptomatic proportion by age* *The size of each circle is proportional to the total number of SARS-CoV-2 positive persons reported in each age group in individual studies, with larger circles indicating a larger sample size.

The following study characteristics were considered when estimating the proportion of asymptomatic infections: study period, study population, country, SARS-CoV-2 infection definition, asymptomatic case definition and follow-up period.

### 2.2. Search Strategy

We searched PubMed, Embase, medRxiv and Google Scholar on 10 September 2020 and 1 March 2021 using keywords COVID-19, SARS-CoV-2, 2019-nCoV, coronavirus disease 2019 AND asymptomatic. We only included studied conducted during January to December 2020, prior to routine vaccination against COVID-19 and the emergence of the alpha, delta or omicron variants. Additionally, this review did not identify or include any studies from regions with the beta variant, which was first detected in South Africa in September 2020. Specific search terms suitable to the individual databases were developed. These search terms included combinations of Medical Subject Headings (MeSH)/Emtree and text words contained in the title and abstract.

### 2.3. Selection Criteria

The article selection process occurred in two phases: 1) title and abstract screen: titles and abstracts of articles identified from the electronic databases and from Internet searches were reviewed; 2) full text review: the full text of articles selected at the title and abstract screen were obtained and reviewed for eligibility. The screening process was completed according to a predefined protocol (Supplementary Document 1). We included all studies reporting

- Proportion of asymptomatic persons among all SARS-CoV-2 infected persons. The numerator includes all SARS-CoV-2 positive persons who were asymptomatic. The denominator includes all SARS-CoV-2 positive persons who tested positive.
- Prevalence of asymptomatic SARS-CoV-2 positive persons among the defined general population. The numerator includes all SARS-CoV-2 positive persons who were asymptomatic. The denominator is the defined study population who were tested for SARS-CoV-2 (e.g., general population in the local community, healthcare workers, patients on hospital admission, nursing home residents).
- Asymptomatic infection: a person with confirmed SARS-CoV-2 infection, who has no symptoms at the time of screening (including the first clinical assessment or laboratory test) and had no symptoms throughout the follow-up period.

We excluded

- Studies published in languages other than English.
- Comments, letters, editorials, case reports, consensus reports and reviews.
- Studies that did not report any age information (e.g., mean or median age) for asymptomatic infections.
- Studies that clearly stated that the SARS-CoV-2 infected persons were included without any follow up and did not distinguish between asymptomatic and pre-symptomatic infections.
- Studies that only tested and enrolled asymptomatic persons and/or mild cases.
- Case studies, case reports and case series with fewer than 20 SARS-CoV-2 infected persons.
- Case studies, case reports and case series that identified SARS-CoV-2 positive persons through contact tracing where only symptomatic persons were tested.
- Serology studies that did not check history of symptoms compatible with SARS-CoV-2 infection and enrolled cases confirmed with SARS-CoV-2 infection by use of IgM only.

### 2.4. Data Extraction

We did not assess study quality because the critical appraisal tools which we planned to use are research design-specific, preclude comparison of the quality of different study designs, and cannot reflect heterogeneity of studies reporting proportions with asymptomatic infections. We did, however, consider several methodological factors in the inclusion/exclusion criteria such as the follow-up period and case identification method. Eight authors (BW, PA, SE, HM, ZL, AT, CB, SG) used an online form in Covidence or a Microsoft Excel spreadsheet to extract the following information: study design, setting, study period, study population (sample size, mean or median age, case definition, etc.), country, follow-up duration, and outcomes (number of people sampled/tested, total number of SARS-CoV-2 positive persons, number of asymptomatic SARS-CoV-2 positive persons).

### 2.5. Statistical Analysis

Most studies reporting prevalence of asymptomatic persons among the tested population were cross-sectional community screening studies without regularly follow-up of SARS-CoV-2 positive persons. We excluded screening studies which clearly stated in the Methods or Discussion sections that they did not follow up any cases or could not distinguish between asymptomatic and pre-symptomatic cases. The number of screening studies, which were eligible and included in our review, was small (n=3). Therefore, the percentage of asymptomatic cases among the tested population was not assessed in the meta-analysis. We only assessed percentage of asymptomatic infections among the confirmed population with laboratory confirmed/clinically diagnosed SARS-CoV-2 infections based on history of exposure to SARS-CoV-2 infection and/or suggestive clinical symptoms of pneumonia (Figure 1).

We used a published method to assess the effect of age on proportion of asymptomatic SARS-CoV-2 positive persons.^21,22^ We anticipated the relationship between the asymptomatic proportion and age in years to be non-linear based on previous reviews.^21,22^ A multilevel mixed-effects logistic regression (QR decomposition) model with a restricted cubic spline was performed to model the asymptomatic proportion as a function of in years. The restricted cubic spline with five knots placed at the ages of 1.4, 13.5, 33.3, 54.5 and 80.4 years was applied, based on Harrell’s recommended percentiles.^23^ Studies were nested within region/country as nested random effects. The model allows for multi-levels of nested clusters of random effects on the assumption that observations within the same cluster are correlated. The outcome measure was the number of asymptomatic persons observed in the study population recorded in binomial form, with the number of SARS-CoV-2 positive persons in the study population as the denominator. When the mean age was not available, we estimated the mean age for each age group using the midpoint of the age band. Any studies reporting the proportion of asymptomatic infections using an age band wider than 20 years were excluded from the meta-analysis. Additional efforts were made to contact the first authors for more information where needed. All analyses were performed using Stata 16.1.^24^

## 3. Results

There were 114 eligible studies. Since a few narrative review and subgroup analyses by age group (e.g., children, adults) have previously been published,^5,7,11,13,16^ only the results of the meta-analysis are discussed here (Figure 1).

### 3.1. Study Characteristics

A total of 38 studies involving 14850 persons were included in the meta-analysis (Table 1) including 13 paediatric studies (n=2729), 8 studies with adults only (n=1156), and 17 studies with children and adults (n=10965), with an age range of 0 to 100 years. Gender was reported in 37 studies including 8931 (61.2%) males and 5574 (38.4%) females. Of 14850 SARS-CoV-2 positive persons included in the meta-analysis, 5498 were from China, 3643 from India, 1519 from Saudi Arabia, 1255 from Bangladesh, 539 from the US, 417 from Kuwait, 230 from Croatia, 220 from Nepal, 213 from Italy, 203 from Greece, 1113 from the rest of the world. Study settings ranged from community screening to hospital treatment/isolation. In 27 studies, SARS-CoV-2 positive patients were enrolled and followed up in the hospital setting.^25-51^ Only three community/international traveler/repatriation screening studies^52-54^ were included as those studies reported follow-up outcomes. In addition, seven disease surveillance studies were included with follow-up outcomes presented in the publication^55-59^ or correspondence with authors.^30,59^ In total, 14 studies including a retrospective online survey^29,34,36,38,39,43,50,51,55,58-62^ which did not clearly state the follow-up period but presented follow-up outcomes, were included in the meta-analysis. The remaining studies followed up patients during the defined follow-up period or during hospital admission. Most SARS-CoV-2 infections were confirmed by RT-PCR. The proportion with asymptomatic infection ranged from 0 to 91.0% with an overall proportion of 44.1%.

**Table 1.**
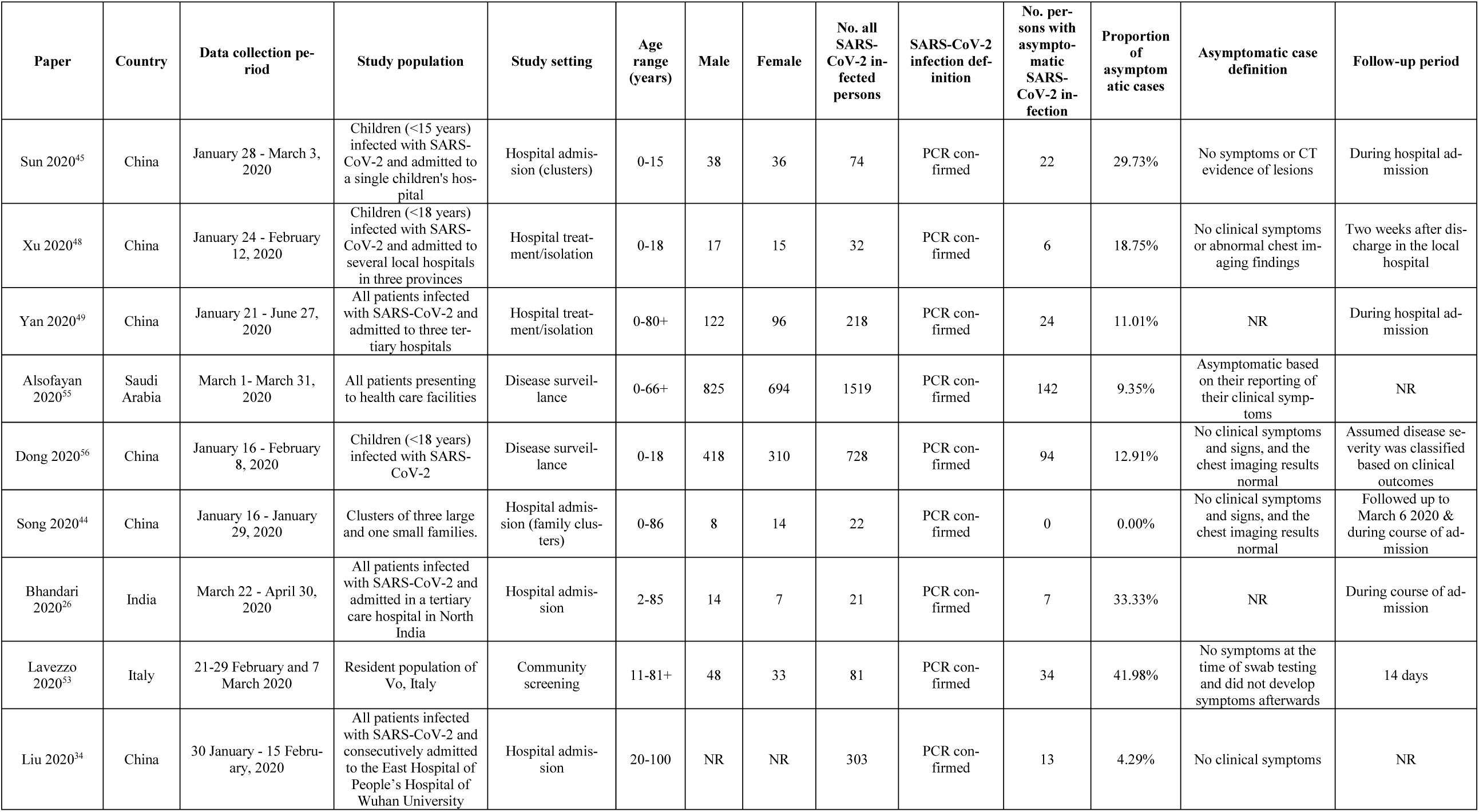
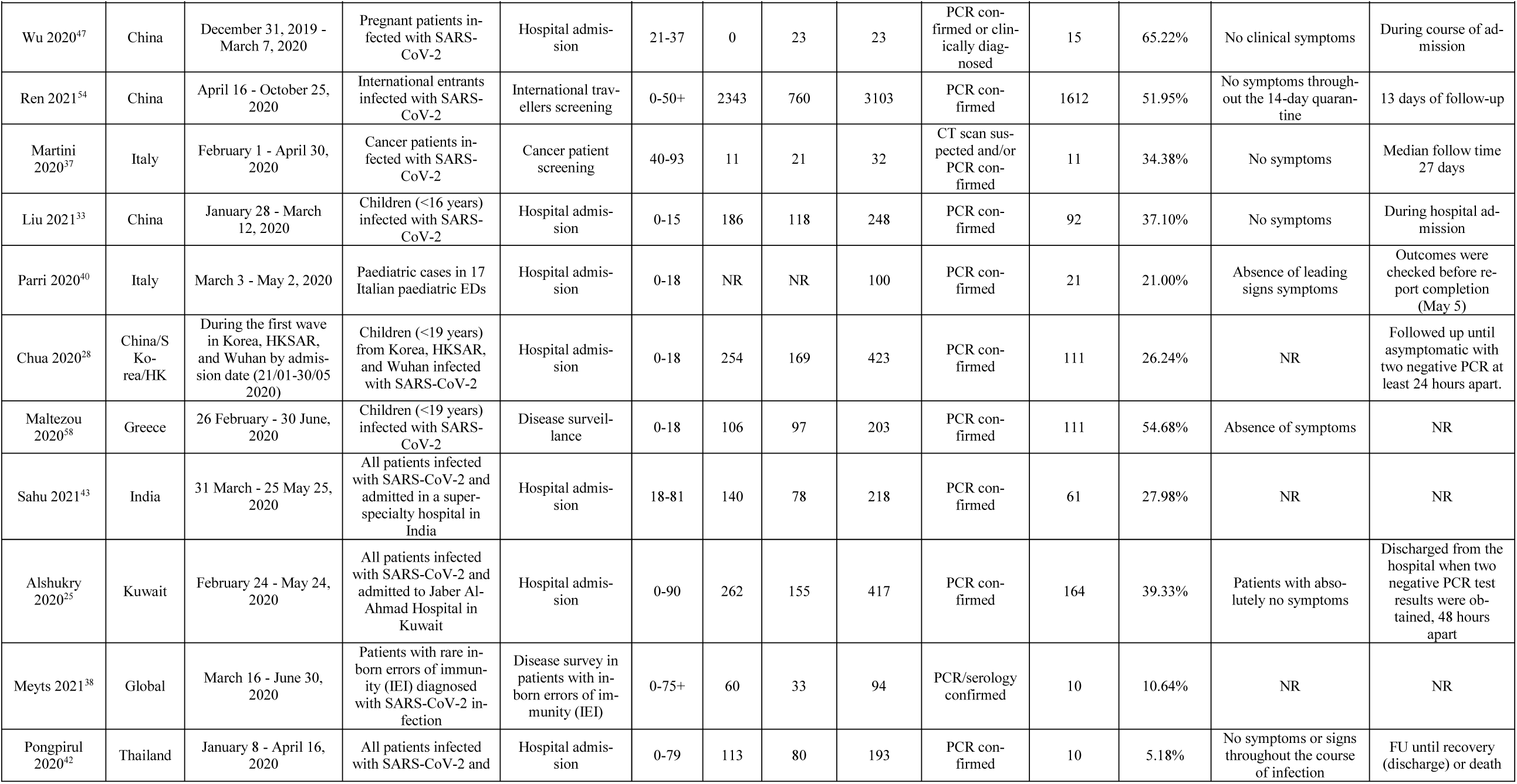
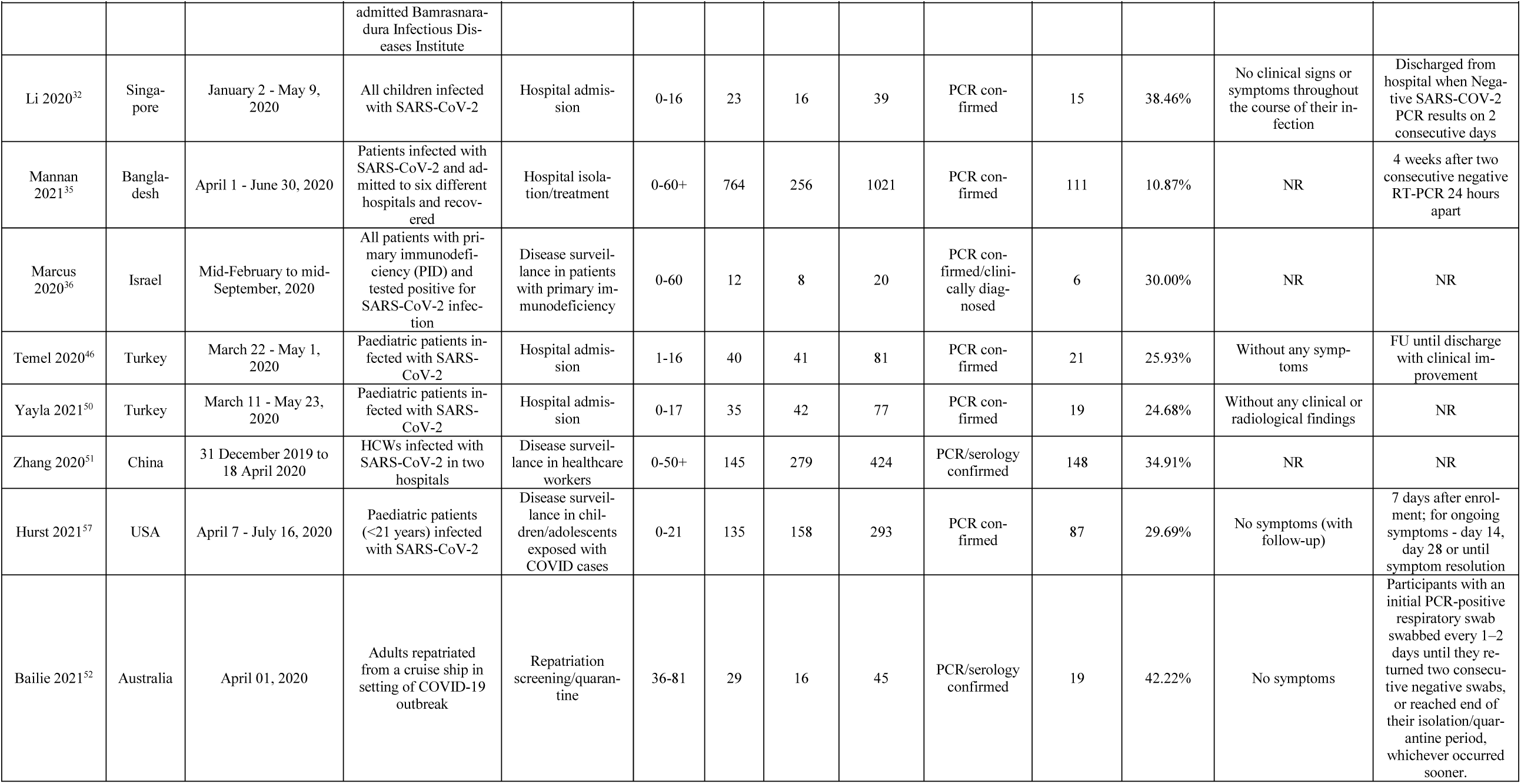
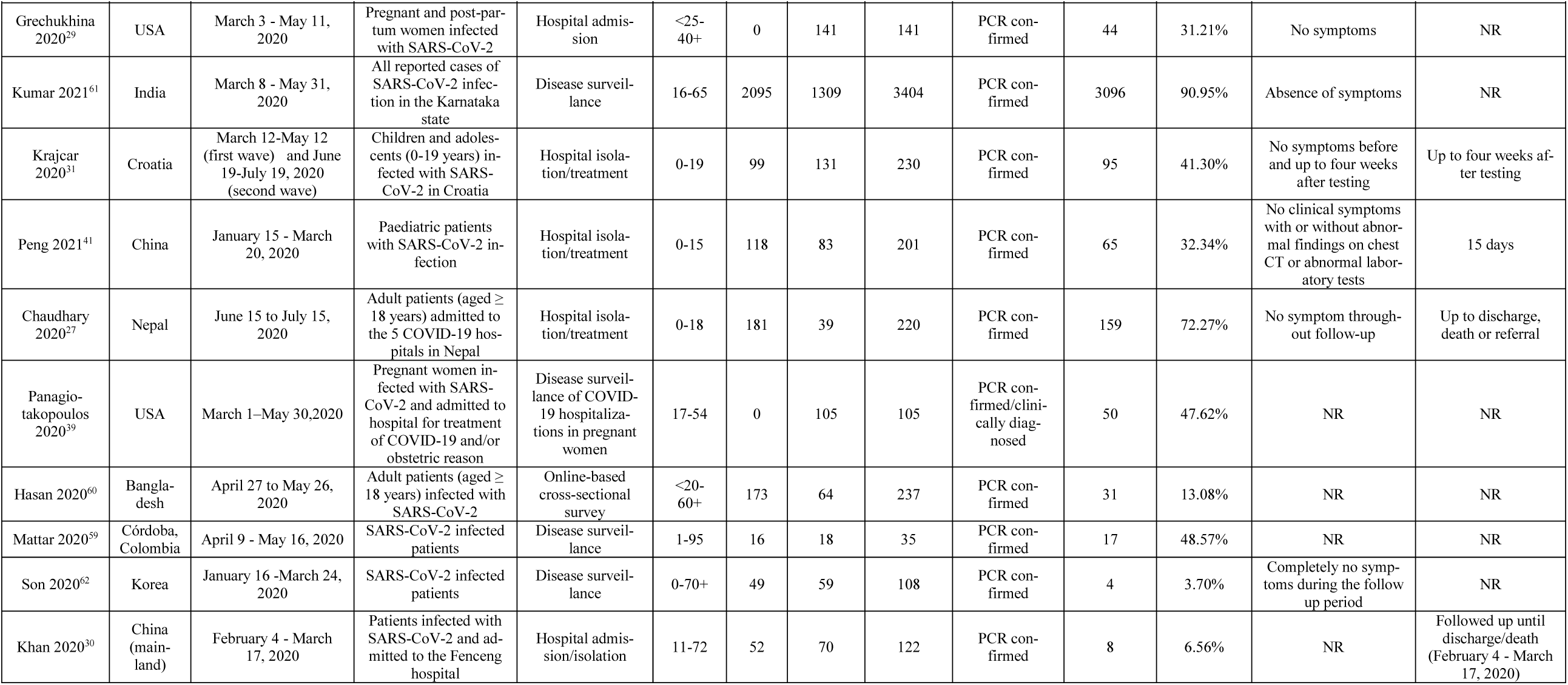
Characteristics of the studies in the meta-analysis

### 3.2. Meta-Analysis

In total, 6556 out of 14850 persons (44.1%, 95%CI 43.3%-45.0%) were reported as asymptomatic throughout the course of infection. The asymptomatic proportion peaked (36.2%, 95%CI: 26.0%-46.5%) at 13.5 years of age, then gradually decreased, levelling out in adults aged 40-50 years, before dropping to 8.1% (95%CI: 3.4%-12.7%) by 90.5 years.

## 4. Discussion

COVID-19 vaccines are highly effective at preventing severe illness, hospitalizations, and death, and have been critical for control the COVID-19 pandemic to restore normal social and economic life.^63^ The COVID-19 vaccine rollout has been extended to children from 5 years for age in many countries including the US, Australia and Europe. Children and young people have been frequently infected with the Delta or Omicron variants due to high transmissibility and as they remain an under-vaccinated group. Previous reviews and meta-analyses^5,7,11,13,16^ have demonstrated that children have the highest proportion of asymptomatic infections, which may jeopardize efforts to prevent transmission within a community. However, these reviews only investigated the proportion with asymptomatic infection across wide age ranges such as adults and children. Our review and meta-analysis is the first to calculate more granular estimates of asymptomatic SARS-CoV-2 infection across the age range.

We found a high proportion of asymptomatic infections in children and young people consistent with previous reviews, which reported the highest proportions in children, and lower in adults, especially older adults.^5,7,11,13,16^ A recent review estimated the pooled percentage of asymptomatic infection to be 41%,^11^ which is similar to our study result of 44%. Four other reviews reported that at least one third of SARS-CoV-2 infections were asymptomatic.^4,12,15,18^ Systematic reviews which were conducted during the early stages of the COVID-19 pandemic,^5-7,9,10,17,19^ however, found that the proportion with asymptomatic infection was much lower than our estimates (13-24%), except for one review^13^ reporting a pooled percentage of asymptomatic cases of 48%. This review^13^ included 16 studies in total and four of them^64-67^ purposefully selected and enrolled asymptomatic SARS-CoV-2 infected persons with 100% of asymptomatic infections. Another review also found that the reported proportion of asymptomatic infections was lower before February 2020 (10%) than after (34%).^16^ This may be due to changes in testing practices, mitigation measures, and dynamics of different circulating variants over time.

Although we attempted to use best available evidence, high heterogeneities were observed in studies. The ideal study design would be a longitudinal study of population, randomly selected to ensure sample was representative and true asymptomatic infections were captured with a well-defined follow up period. However, testing and isolation policies, study settings, follow-up period, and definition of SARS-CoV-2 infection and asymptomatic cases varied between studies. The study settings ranged from hospital admission to universal screening. COVID-19 disease control policies varied between countries. In some countries such as China and Korea, the majority of infected individuals were hospitalised for treatment or isolation regardless of being symptomatic or asymptomatic. In most other countries, asymptomatic cases have only been required to isolate at home. In studies involving hospital admission, the case notes of hospitalised patients were retrospectively reviewed and proportions of asymptomatic cases reported. Almost two thirds of studies included in the review were studies involving hospitalised patients. Consequently, the proportion of asymptomatic infections may be underestimated if symptomatic patients were more likely to be admitted to hospital. Only a small number of community screening studies were identified in our review, mainly because they were cross-sectional and did not follow up asymptomatic individuals. In the inpatient/outpatient screening studies, patients were admitted to hospital for non-COVID-19 conditions such as obstetric admission, dialysis, and elective surgery, which may not represent the broader population in terms of infection and transmission risk. Additionally, asymptomatic infection rates may be overestimated in COVID-19 testing clinics and outbreak settings such as passengers of cruise ships and airplanes.

The follow up period varied significantly between studies. Each publication was meticulously scrutinized and studies were excluded where asymptomatic infections could not adequately be distinguished from pre-symptomatic infections. In the hospital admission studies, cases were frequently followed up. Duration of follow-up varied from a predefined period of days after test positivity or until two negative PCR tests at least 24 hours apart. Some studies, however, did not specify the duration of follow-up in the publication.

The definition of SARS-CoV-2 and asymptomatic infections used were inconsistent between studies. At the beginning of the pandemic, only respiratory symptoms were considered, and loss of taste or smell was not recognized. Some studies defined SARS-CoV-2 infections using clinical diagnostic criteria including radiology findings without a positive PCR/serology test. Although the majority of patients included in our meta-analysis were diagnosed on PCR testing, a small number of patients were diagnosed based on history of exposure to SARS-CoV-2 infection and/or suggestive clinical symptoms of pneumonia but without a positive PCR test.

Since the onset of the SARS-CoV-2 pandemic, multiple new variants of concern have emerged, including the Alpha (B.1.1.7), Beta (B.1.351), Gamma (P.1), Delta (B.1.617.2), and Omicron (B.1.1.529 and BA.2). In December 2020, the UK was first country to start the COVID-19 vaccine roll-out followed by other countries around the world. The Delta variant first detected in India in October 2020 and became the dominant variant globally until the Omicron variant emerged and spread rapidly across the world in November 2021. The population immunity gained through a combination of infection and vaccination has increased over time. Both variants are transmissible than previously circulating strains and Delta has been shown to cause more severe disease in adults compared to Alpha.^68^ It is difficult to determine whether Omicron intrinsically causes milder disease than previous variants of concern. The proportion of asymptomatic infections with Omicron is estimated to be much higher,^69^ which may facilitate more rapid transmission in addition to the variant’s ability to invade both natural and vaccine-induced immunity.^70^ Our review included studies published before 1 March 2021 and only included from pre-vaccine studies era. The proportion of asymptomatic infections reported in our review does not reflect current epidemiological features of the Delta or Omicron variant. The epidemiology of early variants is different to Omicron.

Mass vaccination against COVID-19 with current available vaccines has been highly effective in preventing severe disease and deaths, and reducing healthcare costs and burden.^71-73^ One modelling study demonstrated that the benefits of any COVID-19 vaccine, whether highly, moderately, or modestly efficacious by any trial-defined outcome, would depend on how swiftly and broadly it was implemented.^74^ Given the limited short-term protection provided by current mRNA vaccines against infection nor transmission with the Omicron variant, the priority must be to protect the most vulnerable, especially older adults and those with underlying comorbidities, by ensuring maximum vaccine uptake, boosters as needed and evidence-based mitigations.

Our finding of higher proportion of asymptomatic infections at younger ages suggests that we need to continue to monitor this group closely, especially given that there is currently no vaccine for children under 5 years of age and vaccine uptake is lower in adolescents and children aged 5 years and over compared to adults. An online survey found a higher proportion of parents refused vaccination for their child than the proportion of adults who refused COVID-19 vaccination for themselves.^75^

Since current mRNA vaccines do not prevent infection or transmission with the Omicron variant, further studies are needed on variant-specific mRNA vaccines and other vaccine platforms. Vaccines with a potential to prevent infection in addition to severe COVID-19 could help reduce community transmission and provide population protection as with the nasal influenza vaccines for children.

## Data Availability

All data produced in the present study are available upon reasonable request to the authors.

## Supplementary Materials

Review Protocol.

## Author Contributions

Conceptualization, S.L. and H.S.M.; literature search and review, B.W., P.A., S.E., H.M, Z.A., C.B., A.T. and S.G.; data extraction, B.W., P.A., S.E., H.M., Z.A., A.T., and C.B.; data analysis, B.W.; writing—original draft preparation, B.W.; writing—review and editing, B.W., P.A., S.E., H.M, Z.A., C.B., A.T., S.G., S.L., and H.S.M.. All authors have read and agreed to the published version of the manuscript.

## Institutional Review Board Statement

Not applicable.

## Informed Consent Statement

Not applicable.

## Data Availability Statement

All data are accessible on electronic databases (PubMed, GoogleScholar, Scopus, Embase, Medline, Web of Science).

## Acknowledgments

H.S.M. is supported by a National Health and Medical Research Council (NHMRC) Practitioner Fellowship. The authors would like to express their appreciation to Ms. Natalie Dempster for her generous support and assistance with the literature search. Authors would also like to thank Professor Chang-Hoon Kim at Busan Center for infectious Disease Control and Prevention, and Dr Guang Han at Hubei Cancer Hospital for kindly providing additional information.

